## Supplemental File 1 for "Relevance of Mediterranean diet as a nutritional strategy in diminishing COVID-19 risk: A systematic review"

**S1 File. Detailed search strategies and MeSH.**

**PubMed search strategy**

Keyword: "mediterranean diet and covid"

No filter used.

**ProQuest search strategy**

Keyword: "abstract(Mediterranean diet) AND abstract(Covid)"

Filter used:

- Full text

**Google Scholar search strategy**

Keyword: "allintitle: mediterranean diet covid"

No filter used.

**MeSH for PubMed**

**COVID-19**

A viral disorder generally characterized by high FEVER; COUGH; DYSPNEA; CHILLS; PERSISTENT TREMOR; MUSCLE PAIN; HEADACHE; SORE THROAT; a new loss of taste and/or smell (see AGEUSIA and ANOSMIA) and other symptoms of a VIRAL PNEUMONIA. In severe cases, a myriad of coagulopathy associated symptoms often correlating with COVID-19 severity is seen (e.g., BLOOD COAGULATION; THROMBOSIS; ACUTE RESPIRATORY DISTRESS SYNDROME; SEIZURES; HEART ATTACK; STROKE; multiple CEREBRAL INFARCTIONS; KIDNEY FAILURE; catastrophic ANTIPHOSPHOLIPID ANTIBODY SYNDROME and/or DISSEMINATED INTRAVASCULAR COAGULATION). In younger patients, rare inflammatory syndromes are sometimes associated with COVID-19 (e.g., atypical KAWASAKI SYNDROME; TOXIC SHOCK SYNDROME; pediatric multisystem inflammatory disease; and CYTOKINE STORM SYNDROME). A coronavirus, SARS-CoV-2, in the genus BETACORONAVIRUS is the causative agent.

- COVID 19
- 2019-nCoV Infection
- 2019 nCoV Infection
- 2019-nCoV Infections
- Infection, 2019-nCoV
- SARS-CoV-2 Infection
- Infection, SARS-CoV-2
- SARS CoV 2 Infection
- SARS-CoV-2 Infections
- 2019 Novel Coronavirus Disease
- 2019 Novel Coronavirus Infection
- COVID-19 Virus Infection
- COVID 19 Virus Infection
- COVID-19 Virus Infections
- Infection, COVID-19 Virus
- Virus Infection, COVID-19
- COVID19
- Coronavirus Disease 2019
- Disease 2019, Coronavirus
- Coronavirus Disease-19
- Coronavirus Disease 19
- Severe Acute Respiratory Syndrome Coronavirus 2 Infection
- COVID-19 Virus Disease
- COVID 19 Virus Disease
- COVID-19 Virus Diseases
- Disease, COVID-19 Virus
- Virus Disease, COVID-19
- SARS Coronavirus 2 Infection
- 2019-nCoV Disease
- 2019 nCoV Disease
- 2019-nCoV Diseases
- Disease, 2019-nCoV
- COVID-19 Pandemic
- COVID 19 Pandemic
- Pandemic, COVID-19
- COVID-19 Pandemic

**Mediterranean Diet**

A diet typical of the Mediterranean region characterized by a pattern high in fruits and vegetables, EDIBLE GRAIN and bread, potatoes, poultry, beans, nuts, olive oil and fish while low in red meat and dairy and moderate in alcohol consumption.

- Mediterranean Diet
- Diets, Mediterranean
- Mediterranean Diets
