## Supplemental File 2 for "Relevance of Mediterranean diet as a nutritional strategy in diminishing COVID-19 risk: A systematic review"

| **Risk of bias criteria proposed** | **Yes** | **No** |
| --- | --- | --- |
| Assessment directed at a specific synthesis (e.g., meta-analysis) | | |
| Evidence of funnel plot asymmetry (based on visual inspection of funnel plot or statistical test for funnel plot asymmetry) | N/A | N/A |
| Smaller studies tend to demonstrate more favorable results (based on visual assessment, without funnel plot) | N/A | N/A |
| Clinical decision would differ for estimates from a fixed-effect versus a random-effects model because the findings from a fixed-effect model are closer to the null | N/A | N/A |
| Substantial heterogeneity in the meta-analysis cannot be explained by some clinical or methodological factor | N/A | N/A |
| At least one study is affected by non-publication or non-accessibility |  | ✓ |
| Presence of small (often ‘positive’) studies with for-profit interest in the synthesis |  | ✓ |
| Presence of early studies (ie, set of small, ‘positive’ trials addressing a novel therapy) in the synthesis |  | ✓ |
| Discrepancy in findings between published and unpublished trials |  | ✓ |
| Search strategies were not comprehensive |  | ✓ |
| Methods to identify all available evidence were not comprehensive |  | ✓ |
| Grey literature was not searched | ✓ |  |
| Restrictions to study selection on the basis of language were applied | ✓ |  |
| Industry influence may apply to studies included in the synthesis |  | ✓ |

**S2 File. Reporting bias risk assessment**

**Risk assessment of reporting bias due to selective publication based on Page et al. [16]**

**Risk assessment of reporting bias due to selective non-reporting based on Page et al. [16]**

| **Risk of bias criteria** | **Yes** | **No** |
| --- | --- | --- |
| Assessment directed at study as a whole | | |
| One or more outcomes of interest were clearly measured, but no results were reported |  | ✓ |
| One or more outcomes of interest were reported incompletely so that they could not be entered in a meta-analysis |  | ✓ |
| The study report fails to include results for a key outcome that would be expected to have been reported for such a study |  | ✓ |
| Assessment directed at a specific outcome | | |
| Particular outcome clearly measured but no results were reported |  | ✓ |
| Particular outcome of interest is reported incompletely so that it cannot be entered in a meta-analysis (typically stating only that P>0.05) |  | ✓ |
| Judgement says particular outcome is likely to have been measured and analyzed but not reported on the basis of its results |  | ✓ |
| Composite outcomes are presented without the individual component outcomes |  | ✓ |
| Result reported globally across all groups | ✓ |  |
| Result reported for some groups only |  | ✓ |
| Data were not reported consistently for the outcome of interest |  | ✓ |
| Assessment directed at a specific synthesis | | |
| Selective non-reporting suspected in a number of included studies |  | ✓ |

**Risk assessment of reporting bias due to selection of the reported result based on Page et al. [16]**

| **Risk of bias criteria** | **Yes** | **No** |
| --- | --- | --- |
| Assessment directed at study as a whole | | |
| One or more reported outcomes were not prespecified (unless clear justification for their reporting is provided, such as an unexpected adverse event) |  | ✓ |
| One or more outcomes were reported using measurements, analysis methods or subsets of the data (e.g., subscales) that were not prespecified |  | ✓ |
| One or more retrospective, unplanned, subgroup analyses were reported |  | ✓ |
| Any analyses that had not been planned at the outset of the study were not clearly indicated |  | ✓ |
| Assessment directed at a specific outcome/result | | |
| Particular outcome was not prespecified but results were reported |  |  |
| Reported result for a particular outcome is likely to have been selected, on the basis of the findings, from multiple outcome measurements (e.g., scales, definitions, time points) within the outcome domain |  | ✓ |
| Reported result for a particular outcome is likely to have been selected, on the basis of the findings, from multiple analyses of the data |  | ✓ |
| Reported result for a particular outcome is likely to have been selected, on the basis of the findings, from different subgroup |  | ✓ |

**Risk assessment of reporting bias based on RoBANS 2 [17].**

| **Risk of bias criteria** | | **Yes** | **No** |
| --- | --- | --- | --- |
| **Criteria for ‘low’ risk of bias** | If one or more of the following criteria are met. |  |  |
|  | (1) The protocol that previously determined primary and secondary outcomes was described as planned. | ✓ |  |
|  | (2) Although there was no protocol, most of the expected main outcomes were included. | ✓ |  |
| **Criteria for ‘high’ risk of bias** | If one or more of the following criteria are met. |  |  |
|  | (1) Some of the previously determined primary and secondary outcomes were not reported. |  | ✓ |
|  | (2) Reporting was done using a method that was not previously determined. |  | ✓ |
|  | (3) The outcomes that were not previously determined were reported (exception: when a clear explanation for reporting is provided). |  | ✓ |
|  | (4) The expected main outcomes for the respective study were not reported. |  | ✓ |
| **Criteria for ‘unclear’ risk of bias** | If it is unclear whether the risk of bias belongs to ‘low’ or ‘high’ regarding the selective outcome reporting. |  | ✓ |
